# Preexisting Yellow Fever Virus and West Nile Virus Immunity and Pregnancy Outcomes in a Nigerian Cohort with Endemic Flavivirus Exposure

**DOI:** 10.1101/2025.05.30.25328229

**Authors:** Tae-woo Kim, Bobby Brooke Herrera, Beth Chaplin, Kailee Naito- Keoho, Jerry Ogwuche, Atiene S. Sagay, Charlotte Ajeong Chang, Wei-Kung Wang, Phyllis J. Kanki

## Abstract

**Background:** Yellow fever virus (YFV) and West Nile virus (WNV) co-circulate with other arboviruses, including Zika (ZIKV), dengue (DENV), and chikungunya virus (CHIKV), in sub-Saharan Africa. Associations between preexisting YFV and WNV immunity with symptoms and adverse infant outcomes among pregnant women exposed to flaviviruses are unknown.

**Methods:** We retrospectively studied a prospective cohort of pregnant women enrolled between 2019 and 2022 in Jos, Nigeria. Rapid tests identified ZIKV, DENV, and CHIKV IgM/IgG reactivity for enrollment; 216 women underwent Western blot for YFV and WNV IgG. Logistic regression evaluated associations between arboviral seropositivity and maternal symptoms or adverse infant outcomes. Sequential serology of mother-infant pairs estimated the persistence of passively transferred maternal YFV antibodies.

**Findings:** YFV IgG was detected in 50.5% (109/216) and WNV IgG in 5.1% (11/216) of maternal samples. YFV and WNV seropositivity was significantly associated with maternal symptoms (OR = 2.02, 95% CI: 1.35–3.02, P = 0.001), as was YFV seropositivity alone (OR = 1.77, 95% CI: 1.21–2.61, P < 0.004). CHIKV IgM reactivity was significantly associated with abnormal infant outcomes (OR = 2.38, 95% CI: 1.43–4.02, p = 0.001), but not ZIKV and DENV IgM reactivity. Passive maternal YFV IgG waned in infants at a median of 3.1 months (IQR: 1.65-5.35 months) after birth.

**Interpretation:** YFV and WNV seropositivity was associated with maternal symptoms but not with adverse infant outcomes. Rapid waning of maternal YFV IgG highlights infant vulnerability and supports enhanced surveillance and maternal immunization strategies.

## Introduction

Arthropod-borne viruses (arboviruses) such as yellow fever virus (YFV), West Nile virus (WNV), Zika virus (ZIKV), dengue virus (DENV), and chikungunya virus (CHIKV), pose persistent public health threats to maternal and child health globally, particularly in sub-Saharan Africa and tropical regions (1). Infections with ZIKV, DENV, and CHIKV during pregnancy have been linked to adverse pregnancy outcomes, including congenital abnormalities, stillbirth, and preterm birth (2–4). However, the roles of WNV and YFV infections in pregnancy are limited despite their known ability to cause viremia and neuroinvasive disease (5, 6). Prior studies have focused primarily on acute infection (7), with limited data on the impact of prior YFV and WNV exposure in pregnant women (8).

In flavivirus-endemic regions, sequential or concurrent exposures may trigger complex immunologic interactions. Prior DENV or ZIKV immunity has been associated with both protection and antibody-dependent enhancement (ADE) (9), whereby preexisting antibodies enhance infection severity in humans (10, 11). These dynamics are less well understood for YFV and WNV, especially during pregnancy. Prior studies in general populations have explored interactions between YFV and DENV, including findings that YFV vaccination does not increase DENV severity and that prior DENV exposure may reduce YFV disease severity (12). Whether YFV and/or WNV immunity, acquired through natural infection or vaccination, modulates the course of subsequent arboviral infections such as ZIKV, DENV, or CHIKV remains largely unknown, especially in Africa. This knowledge gap is particularly relevant for pregnancy, where immunologic priming could influence symptom severity or vertical transmission risk.

Arboviruses, including ZIKV, DENV, CHIKV, YFV, and WNV, co-circulate in Nigeria, where annual transmission and overlapping mosquito vectors create an environment of recurring outbreaks and co-infections (13). Our research and surveillance efforts in this region have emphasized cross-reactive immunity in DENV and ZIKV infection and pregnancy outcomes associated with ZIKV, DENV, and CHIKV, which are known for causing abnormal infant outcomes and maternal symptoms (3, 14). In contrast, YFV has received comparatively less attention, likely due to assumptions of widespread vaccine-derived immunity and high efficacy of the YFV vaccine (15). In response to repeated yellow fever epidemics, Nigeria incorporated YFV vaccination into its national immunization schedule in 2004, targeting infants at 9 months of age (16). While this has improved early-life protection, maternal exposure remains understudied, and the population-level impact of YFV and WNV immunity in pregnancy is poorly defined (17).

In endemic settings with significant baseline immunity in pregnant women, passive immunity may provide critical neonatal protection against flavivirus infections (18). Maternal WNV and YFV IgG antibodies against can cross the placenta to provide transitory immunity to newborns (7). However, the duration of this protection remains uncertain, and antibody waning occurs within the first few months of life for other flaviviruses (19). Studies on DENV and ZIKV suggest that passively acquired maternal antibodies may decay leaving infants vulnerable to infections before their immune systems fully mature (20). In the mouse model, sub-neutralizing levels of maternally transferred ZIKV antibodies after waning can increase disease severity in secondary DENV infection and cause antibody-dependent enhancement (ADE) in pups (21). Therefore, understanding the kinetics of maternal antibody persistence is essential to determine whether maternal immunity influences susceptibility to (or protection from) flavivirus infections in neonates.

Our study determined the seroprevalence of YFV and WNV in our prospective cohort of pregnant women in Nigeria, adding to our surveillance of ZIKV, DENV, and CHIKV (3, 14). Using this new data, we re-examined associations between maternal arbovirus serostatus with maternal symptoms and adverse infant outcomes, enabling a more complete analysis of circulating arboviruses. Finally, we measured the rate of antibody passive transfer among YFV seropositive mothers and the time to antibody waning among infants. Understanding the potential impact of WNV and YFV immunity during arbovirus infection in pregnancy has crucial implications for maternal and infant health policies, vaccination strategies, and vector control efforts in flavivirus-endemic regions.

## Research in context

### Evidence before this study

We searched PubMed for studies published before December 31, 2024, using the MeSH terms “Yellow Fever Virus” AND “West Nile Virus” AND “Pregnancy” AND “Infant.” We found no population-based studies that assessed the impact of both YFV and WNV on pregnancy and neonatal health. Most available data on ZIKV and DENV demonstrated associations with adverse outcomes such as microcephaly, stillbirth, and preterm birth. For WNV, case reports suggested possible vertical transmission of WNV, with documented cases of microcephaly, intracranial calcifications, and developmental delays in infants born to infected mothers, but large-scale human studies are lacking. For YFV, existing data primarily come from vaccine safety studies, as live-attenuated vaccination is contraindicated during pregnancy due to theoretical risks of fetal infection, and the impact of natural infection is poorly documented. Most studies focused on high-income settings or vaccine-related outcomes, limiting their relevance to West Africa, where YFV and WNV co-circulate with other flaviviruses. Additionally, no prior studies have systematically evaluated YFV and WNV seroprevalence or their associations with maternal and infant outcomes in Nigeria.

### Added value of this study

While most studies from Asia and the Americas focus on outbreaks by a single viral pathogen, our study addresses gaps in arbovirus co-circulation and maternal health in West Africa. Building on our prior research involving ZIKV, DENV, and CHIKV, we incorporated YFV and WNV to provide a broader serological profile of pregnant women in Nigeria. In a region with YFV vaccination and co-circulation of multiple flaviviruses, we found 50.5% preexisting YFV immunity among pregnant women, associated with 1.77 times increased odds of being symptomatic, but not significantly associated with abnormal infant outcomes. Flavivirus cross-reactivity may have impacted the pathogenesis of adverse maternal and/or infant outcomes. Our study also found low (5.1%) WNV exposure in pregnant women, although seroprevalence rates were insufficient to determine an association with abnormal infant outcomes. We also report a median time to waning of passively transferred maternal YFV IgG in infants of 3.1 months. Our findings contribute to a more comprehensive understanding of arbovirus-related pregnancy risks, informing public health strategies, prenatal screening, and future vaccine recommendations in endemic regions.

### Implications of all the available evidence

Our findings affect maternal health policies, arbovirus surveillance strategies, and public health interventions in endemic regions. While we did not detect a significant association between maternal WNV or YFV seropositivity and adverse pregnancy outcomes, we did see increased odds of adverse outcomes for CHIKV-infected mothers. Larger studies on coinfections and sequential infections are needed to inform targeted screening programs for pregnant women in flavivirus-endemic areas. Rapid waning of maternal YFV antibodies in newborns suggests consideration of public health guidelines on YFV vaccination during pregnancy, particularly in regions where YFV exposure risk outweighs theoretical vaccine safety concerns. Future research should focus on larger-scale prospective cohort studies to confirm the epidemiologic trends, assess long-term neurodevelopmental outcomes in infants, and explore potential consequences of flavivirus co-infection during pregnancy.

## Methods

### Study design and participants

This study retrospectively analyzed archived serum samples from a prospective cohort of pregnant women recruited from antenatal clinics at Jos University Teaching Hospital (JUTH) and Our Lady of Apostles (OLA) Hospital in Jos, Nigeria between April 1, 2019 and January 31, 2022, as previously described (3, 14).

A total of 1,006 pregnant women were screened, with 787 symptomatic and 219 asymptomatic participants based on six arboviral symptoms (fever ≥37.5 °C, rash, headache, arthralgia, conjunctivitis, or myalgia). Rapid diagnostic testing for ZIKV, DENV, and CHIKV IgM/IgG using the ChemBio DPP assay identified 312 women with positive IgM/IgG reactivity to any of the three viruses (200 IgM±IgG and 112 IgG only). Of these, 240 women consented to participate in the prospective study. Twenty-four samples were unavailable, resulting in 216 maternal serum samples included in the final analyses for YFV and WNV IgG testing **(Supplementary Figure 1)**.

Maternal arboviral symptom associations were assessed in all 216 women. Infant outcome data were available for 149 mother-infant pairs. Abnormal outcomes included low birth weight and microcephaly (Z-score ≤-2 standard deviations using sex-and age-specific WHO reference populations) (22), preterm birth (<37 weeks gestation at delivery), stillbirth, and congenital anomalies. A subset of 96 mother-infant pairs had infant samples available for testing; 36 infants born to YFV-seropositive mothers were followed longitudinally to estimate the median time to loss of maternal antibodies. Ethical approval for the study was granted by the institutional review boards of Jos University Teaching Hospital (JUTH/DCS/ADM/127/XXVIII/1338) and the Harvard T.H. Chan School of Public Health (IRB18-1258).

### Western blot analysis

Western blot analysis detected IgG antibodies against YFV and WNV in mothers previously identified as IgM-positive for DENV, ZIKV, or CHIKV (**Figure 1C**). Given the potential for cross-reactivity among flaviviruses, previous studies have shown that detecting envelope (E) and nonstructural protein 1 (NS1) may not reliably distinguish past infections (23). To improve specificity, we determined antibodies against pre-membrane (prM) protein, which can distinguish four flavivirus infections (DENV, ZIKV, WNV, and YFV), as diagnostic for the seroprevalence determination (23).

**Figure 1.**
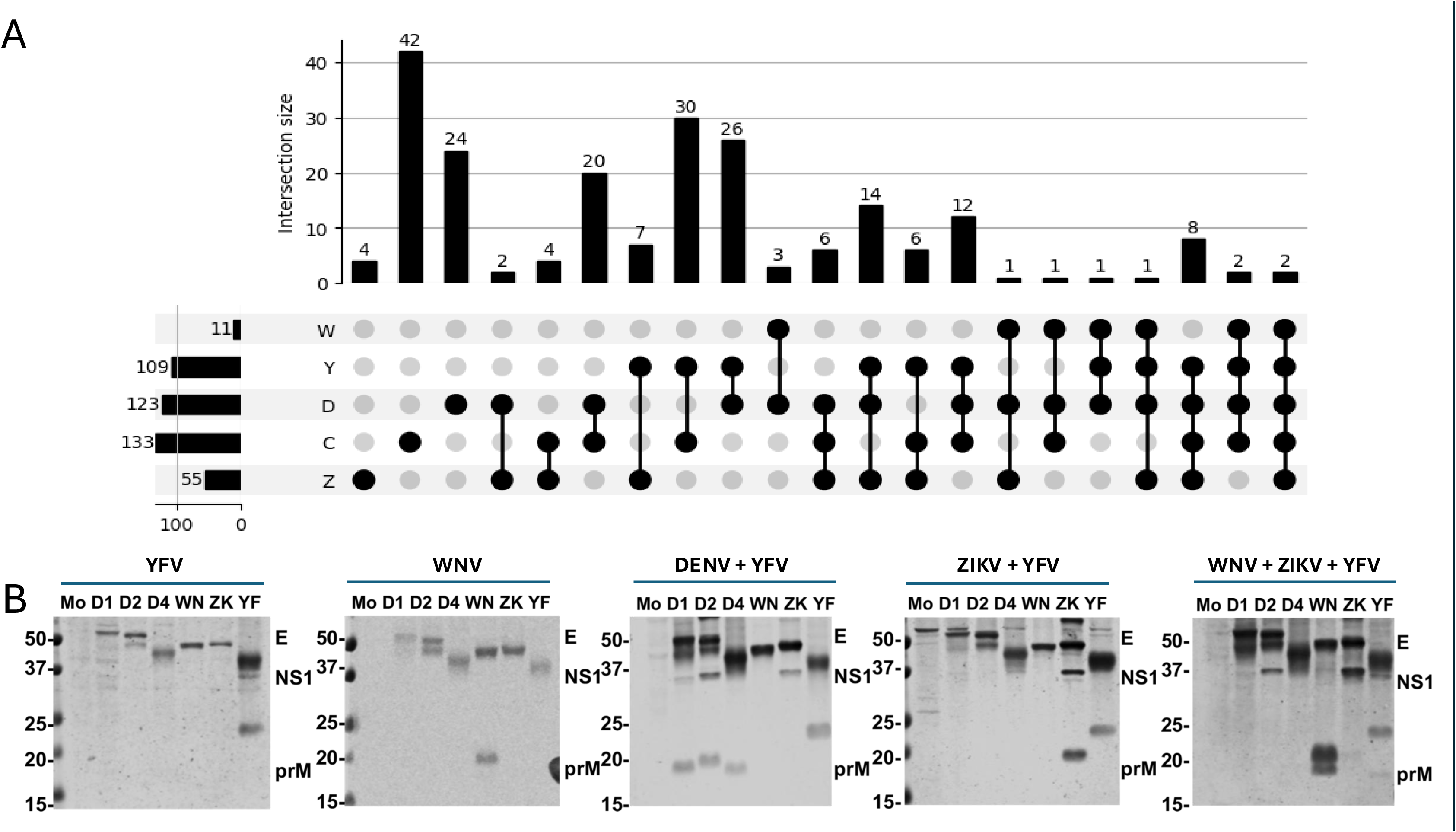
Seroreactivity of Arboviruses Among Pregnant Women in Nigeria (A) UpSet plot displaying the intersections of seroreactivity among ZIKV, CHIKV, DENV, YFV, and WNV. The bar graph represents the number of mothers with seroreactivity of arboviruses, while connected dots indicate virus combinations (B) Venn diagram depicting the overlap of ZIKV, CHIKV, DENV, and WNV seroreactivity. (B) Representative Western blot images for YFV or WNV IgG positivity with other flavivirus in serological testing. The blot includes bands for envelope (E), nonstructural protein 1 (NS1), and pre-membrane (prM) proteins, and the prM band assessed the flavivirus exposure.

Vero cells were infected with YFV or WNV at a multiplicity of infection (MOI) of 1 and incubated at 37 °C until a 50% cytopathic effect was observed. Cells were lysed in NP-40 buffer containing protease and phosphatase inhibitors, incubated on ice for 30 minutes, and clarified by centrifugation (15,200 × g, 10 min, 4 °C).

Protein lysates (100 µL) were resolved on 4–15% SDS-PAGE gels (Bio-Rad, 5671082) and transferred to nitrocellulose membranes. Membranes were blocked in 4% non-fat milk in PBST, cut into strips, and probed overnight at 4 °C with maternal serum (10 µL/sample, diluted in 2% milk/PBST). Blots were washed, incubated with HRP-conjugated anti-human IgG (1:2000, Abcam, Ab97175) for 1 hour at room temperature, and developed with DAB substrate (Thermo-Fisher, 34065). Molecular weight markers (Bio-Rad, 161-0374) were used to identify bands of interest, including E, NS1, and prM proteins. For equivocal or co-reactive samples, membranes were further tested using a half-membrane format containing lysates from six flaviviruses (DENV1, DENV2, DENV4, WNV, ZIKV, and YFV), as described previously (23), to assess cross-reactivity across antigens.

### Data visualization and statistical analysis

Data were managed in FileMaker Pro 18 (Cupertino, CA) and analyzed using STATA/MP 18 (College Station, TX). Descriptive statistics summarized socio-demographic variables and arbovirus seroprevalence.

Associations between maternal arbovirus reactivity and select characteristics (maternal age, trimester, hospital site, year of screening) with clinical outcomes were tested using two-tailed Fisher’s exact tests. Because of low WNV reactivity, WNV and YFV WB reactivity were assessed together, and YFV reactivity was additionally assessed alone for associations. Variables with p-values <0.25 or with known clinical relevance to outcomes were included in multivariable logistic regression to estimate adjusted odds ratios (ORs) and 95% confidence intervals (CIs). Variables with p-values <0.05 in the adjusted logistic regression were considered significant.

Forest plots were generated to display adjusted associations with maternal symptoms and infant outcomes. Venn diagrams and UpSet plots illustrated overlapping IgG reactivities to ZIKV, DENV, YFV, WNV, and CHIKV. Kaplan-Meier survival analysis estimated the time to waning of maternal YFV IgG in infants, with median decay times calculated from the survival curve.

## Results

### Arboviral seroreactivity profiles among pregnant women in Nigeria

Among the 216 women in the final analytical cohort, selected based on IgM and /or IgG reactivity, IgM reactivity was most common for CHIKV (95/216, 44.0%), followed by DENV (83/216, 38.4%), and then ZIKV (34/216, 15.7%). Notably, among IgM-positive people, 3.7% (8/216) showed triple IgM positivity, indicating widespread co-infection. Additionally, 26.9% (58/216) were IgG-positive for at least one of the three viruses despite being IgM-negative, suggesting prior arbovirus exposure.

The western blot analysis confirmed that 109 of 216 individuals (50.5%) were IgG-positive for YFV, while 11 individuals (5.1%) tested positive for WNV. YFV IgG was present across all groups with previous ZIKV, DENV, or CHIKV IgM reactivity, further confirming high levels of flavivirus exposure. Seroreactivity to ZIKV, DENV, CHIKV, WNV, and YFV overlapped significantly based on the IgM and/or IgG results (**Figure 1A**).

### Maternal symptoms and YFV/WNV serostatus

Among 216 women, maternal arboviral symptoms were associated with several serological and demographic variables. YFV/WNV seropositive mothers had significantly higher odds of experiencing symptoms during a subsequent arbovirus infection compared to seronegative mothers (OR = 2.02, 95% CI: 1.35–3.02, P = 0.001, **Table 1**; **Figure 2A**). When YFV reactivity was considered alone, the association remained statistically significant (OR = 1.77, 95% CI: 1.21–2.61, P < 0.004, **Supplementary Table 1**). In contrast, DENV IgM reactivity was inversely associated with symptoms (OR = 0.515, 95% CI: 0.289–0.916, P = 0.024). Neither ZIKV IgM (P=0.484) nor CHIKV IgM (P=0.168) reactivity was significantly associated with symptoms.

**Figure 2.**
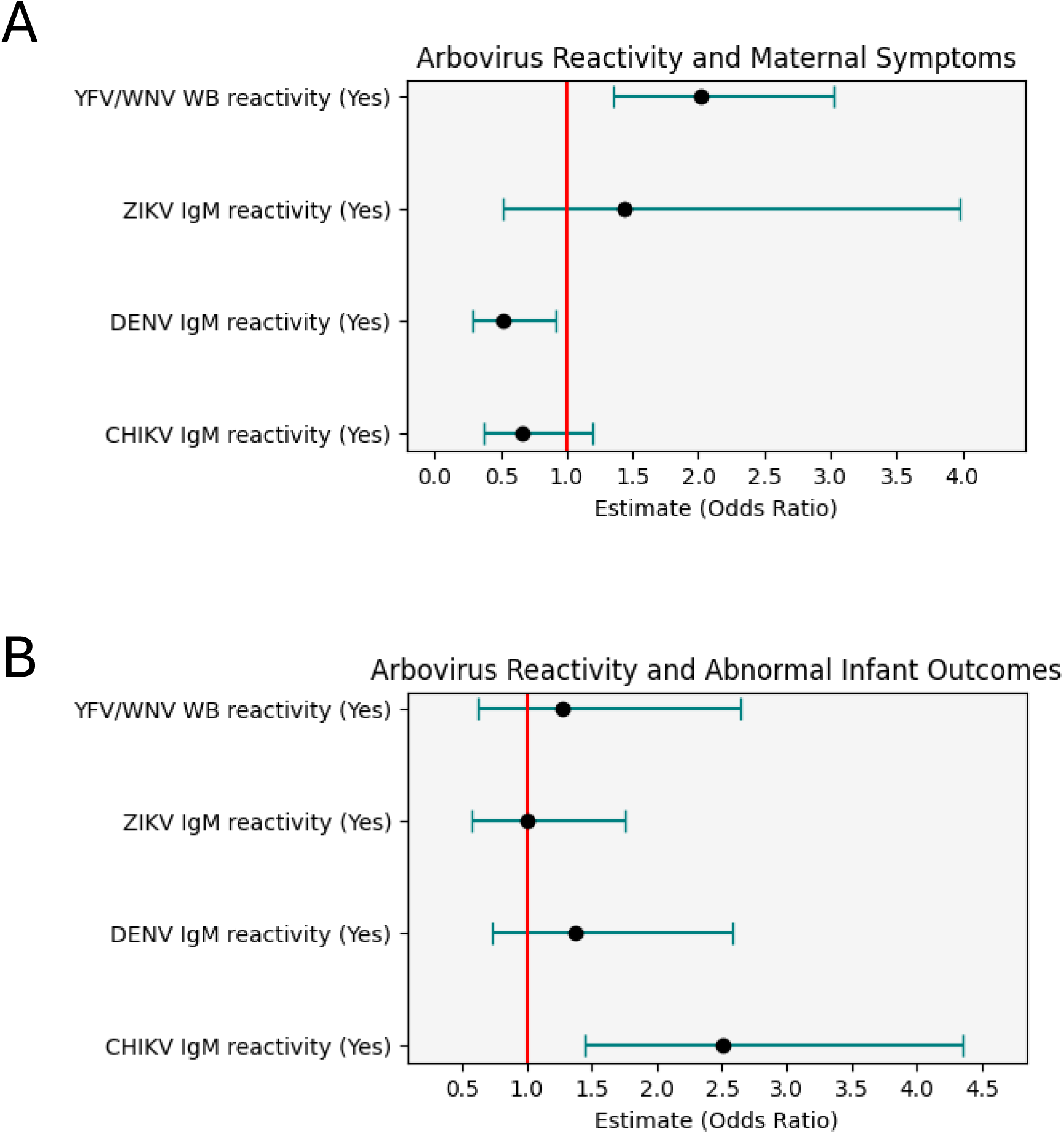
Forest Plot of Arbovirus Seroreactivity and Associations with Maternal Symptoms and Infant Outcomes in Nigerian Cohort (A) Odds ratios (ORs) and 95% confidence intervals (CIs) for the association between arbovirus reactivity and maternal symptoms. (B) Odds ratios (ORs) and 95% confidence intervals (CIs) for the association between arbovirus reactivity and abnormal infant outcomes.

**Table 1.** Associations between YFV and/or WNV seroreactivity and maternal symptoms upon subsequent arbovirus infection among pregnant women in Jos, Nigeria. Statistical analyses include Fisher’s exact test and logistic regression. *Median [interquartile range] is shown for Age (years).

|  | Symptomatic |  | Fisher's exact p-value | Logistic regression |  |
| --- | --- | --- | --- | --- | --- |
|  | No, # (%) | Yes, # (%) |  | Odds ratio (95% CI) | p-value |
| YFV/WNV WB reactivity |  |  | **0.032** |  |  |
| No | 42 (41.2) | 60 (58.8) |  | Ref | - |
| Yes | 31 (27.2) | 83 (72.8) |  | **2.02 (1.35-3.02)** | **0.001** |
| ZIKV IgM reactivity |  |  | 0.430 |  |  |
| No | 64 (35.2) | 118 (64.8) |  | Ref | - |
| Yes | 9 (26.5) | 25 (73.5) |  | 1.44 (0.520-3.98) | 0.484 |
| DENV IgM reactivity |  |  | 0.075 |  |  |
| No | 39 (29.1) | 95 (70.9) |  | Ref | - |
| Yes | 34 (41.5) | 48 (58.5) |  | **0.515 (0.289-0.916)** | **0.024** |
| CHIKV IgM reactivity |  |  | 0.311 |  |  |
| No | 37 (30.6) | 84 (69.4) |  | Ref | - |
| Yes | 36 (37.9) | 59 (62.1) |  | 0.668 (0.376-1.19) | 0.168 |
| Site |  |  | **<0.001** |  |  |
| JUTH | 14 (14.6) | 82 (85.4) |  |  |  |
| OLA | 59 (49.2) | 61 (50.8) |  |  |  |
| Year screened |  |  | 0.220 |  |  |
| 2019 | 26 (28.9) | 64 (71.1) |  | Ref | - |
| 2020 | 28 (41.8) | 39 (58.2) |  | **0.579 (0.430-0.779)** | **<0.001** |
| 2021-22 | 19 (32.2) | 40 (67.8) |  | **0.817 (0.784-0.852)** | **<0.001** |
| Age (years)* | 30 [26-34] |  |  | 0.995 (0.932-1.06) | 0.875 |
| Trimester |  |  | 0.473 |  |  |
| 1st&2nd (1-27 wks) | 42 (36.2) | 74 (63.8) |  |  |  |
| 3rd (≥28 wks) | 31 (31.3) | 68 (68.7) |  |  |  |

Symptom prevalence also varied by study site, with a higher proportion of symptomatic women at JUTH compared to OLA (85.4% vs. 50.8%, P < 0.001). Screening year also impacted symptom odds, with women enrolled in 2020 (OR = 0.579, 95% CI: 0.430–0.779, P < 0.001) and 2021–22 (OR = 0.817, 95% CI: 0.784–0.852, P < 0.001) less likely to report symptoms compared to those screened in 2019, considering environmental and vector exposure differences over time. Neither maternal age nor trimester at screening was significantly associated with symptoms.

### Infant outcomes and maternal arbovirus serostatus

Among 149 mothers with infant outcome data, abnormal infant outcomes were not significantly associated with maternal YFV/WNV IgG serostatus in neither Fisher’s exact test nor multivariable logistic regression (OR = 1.28, P = 0.503, **Table 2**; **Figure 2B**). Similarly, YFV IgG seropositivity alone did not predict infant abnormalities (**Supplementary Table 2**).

**Table 2.** Associations between YFV and/or WNV seroreactivity and abnormal infant outcomes upon subsequent arbovirus infection among pregnant women in Jos, Nigeria. Statistical analyses include Fisher’s exact test and logistic regression. *Median [interquartile range] is shown for Age (years).

|  | Abnormal infant outcome |  | Fisher's exact p-value | Logistic regression |  |
| --- | --- | --- | --- | --- | --- |
|  | No, # (%) | Yes, # (%) |  | Odds ratio (95% CI) | p-value |
| YFV/WNV WB reactivity |  |  | 0.364 |  |  |
| No | 46 (75.4) | 15 (24.6) |  | Ref | - |
| Yes | 60 (68.2) | 28 (31.8) |  | 1.28 (0.621-2.64) | 0.503 |
| ZIKV IgM reactivity |  |  | 1.000 |  |  |
| No | 86 (71.1) | 35 (28.9) |  | Ref | - |
| Yes | 20 (71.4) | 8 (28.6) |  | 1.01 (0.580-1.75) | 0.976 |
| DENV IgM reactivity |  |  | 0.855 |  |  |
| No | 61 (79.1) | 26 (29.9) |  | Ref | - |
| Yes | 45 (72.6) | 17 (27.4) |  | 1.37 (0.733-2.58) | 0.322 |
| CHIKV IgM reactivity |  |  | **0.030** |  |  |
| No | 61 (79.2) | 16 (20.8) |  | Ref | - |
| Yes | 45 (62.5) | 27 (37.5) |  | **2.51 (1.45-4.35)** | **0.001** |
| Site |  |  | 0.567 |  |  |
| JUTH | 32 (68.1) | 15 (31.9) |  |  |  |
| OLA | 74 (72.6) | 28 (27.4) |  |  |  |
| Year screened |  |  | 0.326 |  |  |
| 2019 | 51 (66.2) | 26 (33.8) |  |  |  |
| 2020 | 26 (72.2) | 10 (27.8) |  |  |  |
| 2021-22 | 29 (80.6) | 7 (19.4) |  |  |  |
| Age (years) | 30 [26-33] |  |  | **1.07 (1.02-1.11)** | **0.003** |
| Trimester |  |  | 0.468 |  |  |
| 1st&2nd (1-27 wks) | 60 (74.1) | 21 (25.9) |  |  |  |
| 3rd (≥28 wks) | 46 (67.7) | 22 (32.3) |  |  |  |

However, CHIKV IgM reactivity emerged as a significant predictor. Mothers positive for CHIKV IgM had over two times higher odds of having infants with abnormal outcomes (OR = 2.51, 95% CI: 1.48–4.20, P = 0.001). Maternal age emerged as a significant predictor (OR = 1.07, 95% CI: 1.02–1.11, P = 0.004), suggesting increased risk with advancing age. Trimester and screening year did not significantly affect abnormal infant outcomes.

### Maternal immunity passive transfer

Among 96 infants with available follow-up samples, 51 were born to YFV IgG-seropositive mothers. Of these, 36 infants (70.6%) tested YFV IgG-positive at birth, indicating transplacental transfer of maternal antibodies. The longitudinal follow-up revealed a progressive decline in passively acquired IgG over time, with variable rates of antibody waning across individuals (**Supplementary Figure 3)**. By six months of age, 22 infants (22/26, 84.6%) had waning antibodies. Kaplan-Meier survival analysis estimated a median time to IgG waning of 3.1 months (IQR: 1.65-5.35 months) (**Figure 3**). The results demonstrate that over 50% of infants become YFV seronegative within the first four months, while most maternal antibodies begin to wane over 6-12 months (24). These findings demonstrate the transience of maternally derived YFV immunity in infants and highlight the vulnerability of YFV exposure in early infancy.

**Figure 3.**
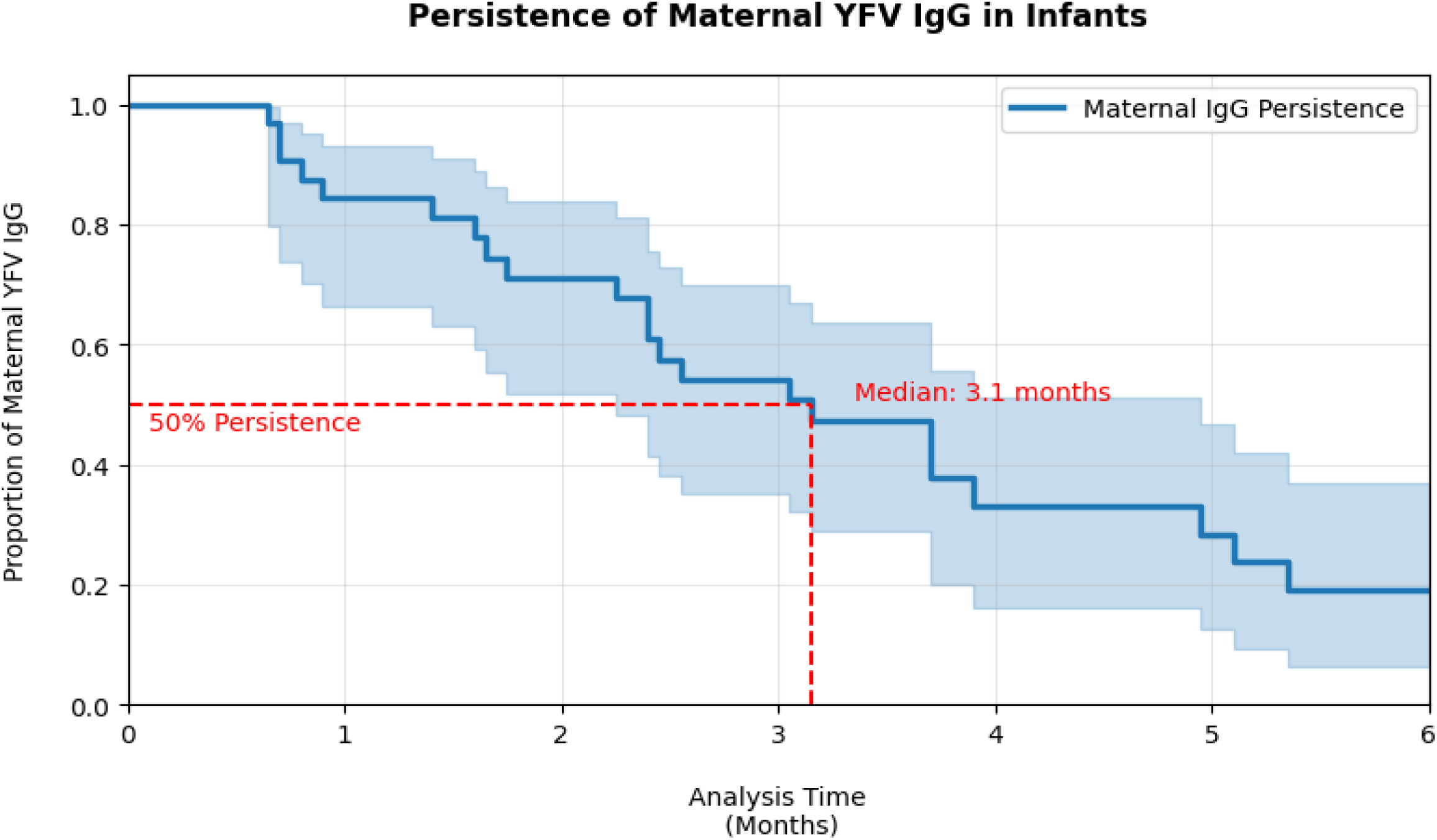
Persistence of Maternal YFV IgG in Infants in North-Central Nigeria. Kaplan-Meier survival curve showing the proportion of infants with detectable maternal YFV IgG over time. The blue line represents the survival estimate, and the shaded area represents the 95% confidence interval. The median time to IgG waning is 3·1 months (IQR: 1·65-5·35 months), as indicated by the red dashed lines.

## Discussion

In this retrospective study of pregnant women and their infants, we observed high seroreactivity to arboviruses, especially YFV (109/216, 50.5%), DENV (124/216, 57.4%), and CHIKV (133/216, 61.5%), and lower seroreactivity to WNV (11/216, 5.1%), based on IgM and/or IgG results (Figure 1A). YFV and WNV IgG seropositivity were significantly associated with increased risk of maternal symptoms during a subsequent arbovirus infection but not with risk of adverse infant outcomes. In a subset analysis of 36 infants, maternal YFV IgG waned with a median duration of 3.1 months (IQR: 1.65-5.35 months) and indicated limited protection of infants after waning since the YFV vaccine is not given until 9 months (25). These findings raise important considerations about the sufficiency of maternally derived immunity in these highly endemic regions.

High IgM reactivity to CHIKV and DENV reflects intense natural transmission, whereas broad YFV IgG seropositivity likely reflects a combination of natural infection and widespread vaccination coverage through Nigeria’s national childhood immunization program, established in 2004 (26). The observed 50.5% YFV seropositivity in our cohort is notably higher than the predicted YFV-17D vaccine coverage (30-40%) in Nigeria up to 2016 (27), likely reflecting the recent surge in epidemics along with vaccine campaigns. In contrast, WNV IgG was detected in only a small subset of individuals (6.2%). Its low prevalence limited the statistical power to assess independent associations with maternal or infant outcomes. Most observed effects were, therefore, likely driven by YFV seroreactivity. These findings highlight a high degree of flavivirus exposure from natural infections and vaccination, particularly for YFV and DENV. Given the endemic nature of these viruses, our results highlight the importance of continued serological surveillance to mitigate the burden of flavivirus-related morbidity among pregnant women in high-risk regions.

The seropositivity to multiple viruses reflected the diagnostic complexity in endemic regions. This is particularly relevant for flaviviruses, which share considerable sequence homology across structural (E, prM) and non-structural (NS1) proteins (28). The live-attenuated YFV vaccine expresses both structural and non-structural proteins (29). As a result, the YFV vaccine-induced broad immune response may be indistinguishable from natural infection using standard serological assays. This cross-reactive immune landscape complicates efforts to distinguish YFV vaccine-derived immunity from natural YFV infection, as well as to interpret recent flavivirus infections in individuals with prior YFV vaccination. More specific diagnostics tools, such as virus-specific neutralization assays and multiplexed viral antigens or nucleic acid-based assays, are needed to improve flavivirus surveillance and attribution of clinical diseases.

YFV and WNV IgG positivity were associated with 2.02 times the odds of maternal symptoms, even after adjusting for confounders. YFV reactivity alone did not reach significance in a univariate model but emerged as significant in multivariable regression. The multivariable regression models suggest possible synergistic effects of co-infection or immune priming from previous flavivirus exposures. These results align with other studies from endemic settings, where past flavivirus immunity exacerbates the severity of clinical symptoms, potentially through weakly neutralizing antibodies or in lower concentrations through Fc-mediated functions (30). The observation raises the possibility that prior YFV or WNV exposure may influence inflammatory or symptomatic responses during pregnancy with other arbovirus coinfections. This is consistent with the prior finding where Japanese encephalitis virus (JEV) neutralizing antibodies were associated with increased symptomatic DENV illness in a pediatric cohort in Thailand (31).

We explored whether YFV/WNV immunity modifies the effect of ZIKV or DENV IgM reactivity on maternal symptoms by testing for the interaction in our multiple logistic regression model. Although the interaction term was not statistically significant (p = 0.129), predictive margins showed that symptom probability was higher among ZIKV/DENV IgM-positive mothers with YFV/WNV IgG (70.9%) compared to those without IgG (47.8%), based on predictive margins from the regression model (data not shown). This suggests a possible differential effect that merits further investigation.

Despite the burden of maternal symptoms, neither YFV/WNV seroreactivity nor IgM reactivity with ZIKV or DENV was associated with adverse infant outcomes in this study. Instead, CHIKV IgM reactivity was robustly associated with abnormal infant outcomes (OR = 2.51). The association brings concerns about the neurotropic potential of CHIKV and its potential role in pregnancy complications (32). However, the association between YFV and WNV with maternal symptoms but not with infant outcomes suggests the limitation of flaviviruses’ vertical transmission in the presence of YFV or WNV prior exposures. Additionally, the rapid waning of maternal YFV IgG in infants raises concerns about a window of vulnerability before vaccine-induced immunity is established. Infants may be unprotected during outbreaks occurring in the first months of life. While considerably higher than WNV, YFV IgG seroprevalence was still only 50.5% indicating that half of mothers have no immunity to transfer to their infants. This suggests the revision of maternal vaccination strategies for women of childbearing age and education on safe mosquito protection and environmental control interventions such as protective clothing, mosquito nets, and safe repellents, at least until 9 months, the age of YFV vaccination, for newborns and new mothers in endemic regions.

Our study has limitations related to the lack of temporal resolution for exposure due to retrospective design. The variability in follow-up timing and incomplete infant outcome data may have reduced the power of associations. We could not distinguish between the YFV vaccine versus natural immunity. Finally, our study population included only pregnant women positive for ZIKV, DENV, and CHIKV IgM and IgG and their infants, which enabled us to look at the role of YFV and WNV immunity in infection with those viruses, but we did not have a negative control group. Future studies will need prospective longitudinal cohort designs with better-defined infection timing, maternal-fetal transmission dynamics, and durability of passive immunity.

Nonetheless, this study provides novel seroprevalence data on YFV and WNV immunity among pregnant women in Nigeria and on the persistence of passively transferred maternal YFV IgG antibodies in neonates. The increased odds of arboviral symptoms among those with YFV and/or WNV antibodies during pregnancy suggest the possibility of ADE in secondary flavivirus infections, which indicates the need for further studies. Understanding how immunity from prior vaccination or infection affects pregnancy outcomes and neonatal vulnerability remains essential for optimizing vaccination policies and clinical care in endemic settings.

## Contributors

Conceptualization: TWK, BBH, WKW, PJK; data curation: TWK, BRC, JO, ASS; Formal analysis: TWK, BRC, CAC, PJK; Investigation: TWK, BRC, KNK, CAC; Funding acquisition: WKW, PJK; project administration: JO, ASS, PJK; writing, review and editing: TWK, BBH, BRC, KNK, JO, ASS, CAC, WKW, PJK.

## Supporting information

Kim_Supplemental

## Data Availability

All data in the present study are available upon reasonable request to the corresponding author

## Acknowledgments

The authors acknowledge our patient participants and clinic staff in Jos, Nigeria for their important participation in this study. We also thank the Motsepe Presidential Research Accelerator Fund for Africa and the National Institutes of Health under award number R21AI137840 (Kanki) and R01AI149502 (Wang) for supporting the research reported in this publication.

## References

1. Guarner J, Hale GL. Four human diseases with significant public health impact caused by mosquito-borne flaviviruses: West Nile, Zika, dengue and yellow fever. Semin Diagn Pathol. 2019;36(3):170–6.

2. Brar R, Sikka P, Suri V, Singh MP, Suri V, Mohindra R, et al. Maternal and fetal outcomes of dengue fever in pregnancy: a large prospective and descriptive observational study. Archives of Gynecology and Obstetrics. 2021;304(1):91–100.

3. Sagay AS, Hsieh SC, Dai YC, Chang CA, Ogwuche J, Ige OO, et al. Chikungunya virus antepartum transmission and abnormal infant outcomes in a cohort of pregnant women in Nigeria. Int J Infect Dis. 2024;139:92–100.

4. Gilbert RK, Petersen LR, Honein MA, Moore CA, Rasmussen SA. Zika virus as a cause of birth defects: Were the teratogenic effects of Zika virus missed for decades? Birth Defects Res. 2023;115(3):265–74.

5. Malik S, Pandey I, Kishore S, Sundarrajan T, Nargund SL, Ghosh A, et al. Yellow fever virus, a mosquito-borne flavivirus posing high public health concerns and imminent threats to travellers - an update. Int J Surg. 2023;109(2):134–7.

6. Santini M, Haberle S, Židovec-Lepej S, Savić V, Kusulja M, Papić N, et al. Severe West Nile Virus Neuroinvasive Disease: Clinical Characteristics, Short-and Long-Term Outcomes. Pathogens. 2022;11(1).

7. Howard-Jones AR, Pham D, Sparks R, Maddocks S, Dwyer DE, Kok J, et al. Arthropod-Borne Flaviviruses in Pregnancy. Microorganisms [Internet]. 2023; 11(2).

8. O’Leary DR, Kuhn S, Kniss KL, Hinckley AF, Rasmussen SA, Pape WJ, et al. Birth outcomes following West Nile Virus infection of pregnant women in the United States: 2003-2004. Pediatrics. 2006;117(3):e537–45.

9. Khandia R, Munjal A, Dhama K, Karthik K, Tiwari R, Malik YS, et al. Modulation of Dengue/Zika Virus Pathogenicity by Antibody-Dependent Enhancement and Strategies to Protect Against Enhancement in Zika Virus Infection. Front Immunol. 2018;9:597.

10. Rodriguez-Barraquer I, Costa F, Nascimento EJM, Nery NJ, Castanha PMS, Sacramento GA, et al. Impact of preexisting dengue immunity on Zika virus emergence in a dengue endemic region. Science. 2019;363(6427):607-10.

11. Zambrana JV, Hasund CM, Aogo RA, Bos S, Arguello S, Gonzalez K, et al. Primary exposure to Zika virus is linked with increased risk of symptomatic dengue virus infection with serotypes 2, 3, and 4, but not 1. Sci Transl Med. 2024;16(749):eadn2199.

12. Luppe MJ, Verro AT, Barbosa AS, Nogueira ML, Undurraga EA, da Silva NS. Yellow fever (YF) vaccination does not increase dengue severity: A retrospective study based on 11,448 dengue notifications in a YF and dengue endemic region. Travel Med Infect Dis. 2019;30:25–31.

13. Shaibu JO, Akinyemi KO, Uzor OH, Audu RA, Bola Oriowo Oyefolu A. Molecular surveillance of arboviruses in Nigeria. BMC Infect Dis. 2023;23(1):538.

14. Ogwuche J, Chang CA, Ige O, Sagay AS, Chaplin B, Kahansim ML, et al. Arbovirus surveillance in pregnant women in north-central Nigeria, 2019-2022. J Clin Virol. 2023;169:105616.

15. Gotuzzo E, Yactayo S, Córdova E. Efficacy and duration of immunity after yellow fever vaccination: systematic review on the need for a booster every 10 years. Am J Trop Med Hyg. 2013;89(3):434–44.

16. Ophori EA, Tula MY, Azih AV, Okojie R, Ikpo PE. Current trends of immunization in Nigeria: prospect and challenges. Trop Med Health. 2014;42(2):67–75.

17. Schnyder JL, de Jong HK, Bache BE, Schaumburg F, Grobusch MP. Long-term immunity following yellow fever vaccination: a systematic review and meta-analysis. Lancet Glob Health. 2024;12(3):e445–e56.

18. Albrecht M, Pagenkemper M, Wiessner C, Spohn M, Lütgehetmann M, Jacobsen H, et al. Infant immunity against viral infections is advanced by the placenta-dependent vertical transfer of maternal antibodies. Vaccine. 2022;40(11):1563–71.

19. Clapham H, Cummings DAT, Nisalak A, Kalayanarooj S, Thaisomboonsuk B, Klungthong C, et al. Epidemiology of Infant Dengue Cases Illuminates Serotype-Specificity in the Interaction between Immunity and Disease, and Changes in Transmission Dynamics. PLOS Neglected Tropical Diseases. 2015;9(12):e0004262.

20. Chau TN, Hieu NT, Anders KL, Wolbers M, Lien le B, Hieu LT, et al. Dengue virus infections and maternal antibody decay in a prospective birth cohort study of Vietnamese infants. J Infect Dis. 2009;200(12):1893–900.

21. Fowler AM, Tang WW, Young MP, Mamidi A, Viramontes KM, McCauley MD, et al. Maternally Acquired Zika Antibodies Enhance Dengue Disease Severity in Mice. Cell Host & Microbe. 2018;24(5):743–50.e5.

22. Organization WH. WHO Child Growth Standards: Length/height-for-age, weight-for-age, weight-for-length, weight-for-height and body mass index-for-age: Methods and development Geneva: World Health Organization; 2006 [http://www.who.int/childgrowth/standards/second_set/technical_report_2.pdf]

23. Chen G-H, Dai Y-C, Hsieh S-C, Tsai J-J, Sy AK, Jiz M, et al. Detection of anti-premembrane antibody as a specific marker of four flavivirus serocomplexes and its application to serosurveillance in endemic regions. Emerging Microbes & Infections. 2024;13(1):2301666.

24. Niewiesk S. Maternal antibodies: clinical significance, mechanism of interference with immune responses, and possible vaccination strategies. Front Immunol. 2014;5:446.

25. Domingo C, Fraissinet J, Ansah PO, Kelly C, Bhat N, Sow SO, et al. Long-term immunity against yellow fever in children vaccinated during infancy: a longitudinal cohort study. Lancet Infect Dis. 2019;19(12):1363–70.

26. Nomhwange T, Jean Baptiste AE, Ezebilo O, Oteri J, Olajide L, Emelife K, et al. The resurgence of yellow fever outbreaks in Nigeria: a 2-year review 2017-2019. BMC Infect Dis. 2021;21(1):1054.

27. Shearer FM, Moyes CL, Pigott DM, Brady OJ, Marinho F, Deshpande A, et al. Global yellow fever vaccination coverage from 1970 to 2016: an adjusted retrospective analysis. Lancet Infect Dis. 2017;17(11):1209–17.

28. da Fonseca NJ, Lima Afonso MQ, Pedersolli NG, de Oliveira LC, Andrade DS, Bleicher L. Sequence, structure and function relationships in flaviviruses as assessed by evolutive aspects of its conserved non-structural protein domains. Biochemical and Biophysical Research Communications. 2017;492(4):565–71.

29. Mateus J, Grifoni A, Voic H, Angelo MA, Phillips E, Mallal S, et al. Identification of Novel Yellow Fever Class II Epitopes in YF-17D Vaccinees. Viruses. 2020;12(11).

30. Pierson TC, Xu Q, Nelson S, Oliphant T, Nybakken GE, Fremont DH, et al. The stoichiometry of antibody-mediated neutralization and enhancement of West Nile virus infection. Cell Host Microbe. 2007;1(2):135–45.

31. Anderson KB, Gibbons RV, Thomas SJ, Rothman AL, Nisalak A, Berkelman RL, et al. Preexisting Japanese encephalitis virus neutralizing antibodies and increased symptomatic dengue illness in a school-based cohort in Thailand. PLoS Negl Trop Dis. 2011;5(10):e1311.

32. Foeller ME, Nosrat C, Krystosik A, Noel T, Gérardin P, Cudjoe N, et al. Chikungunya infection in pregnancy - reassuring maternal and perinatal outcomes: a retrospective observational study. Bjog. 2021;128(6):1077–86.

