## Supplementary material for "Preexisting Yellow Fever Virus and West Nile Virus Immunity and Pregnancy Outcomes in a Nigerian Cohort with Endemic Flavivirus Exposure": Kim_Supplemental

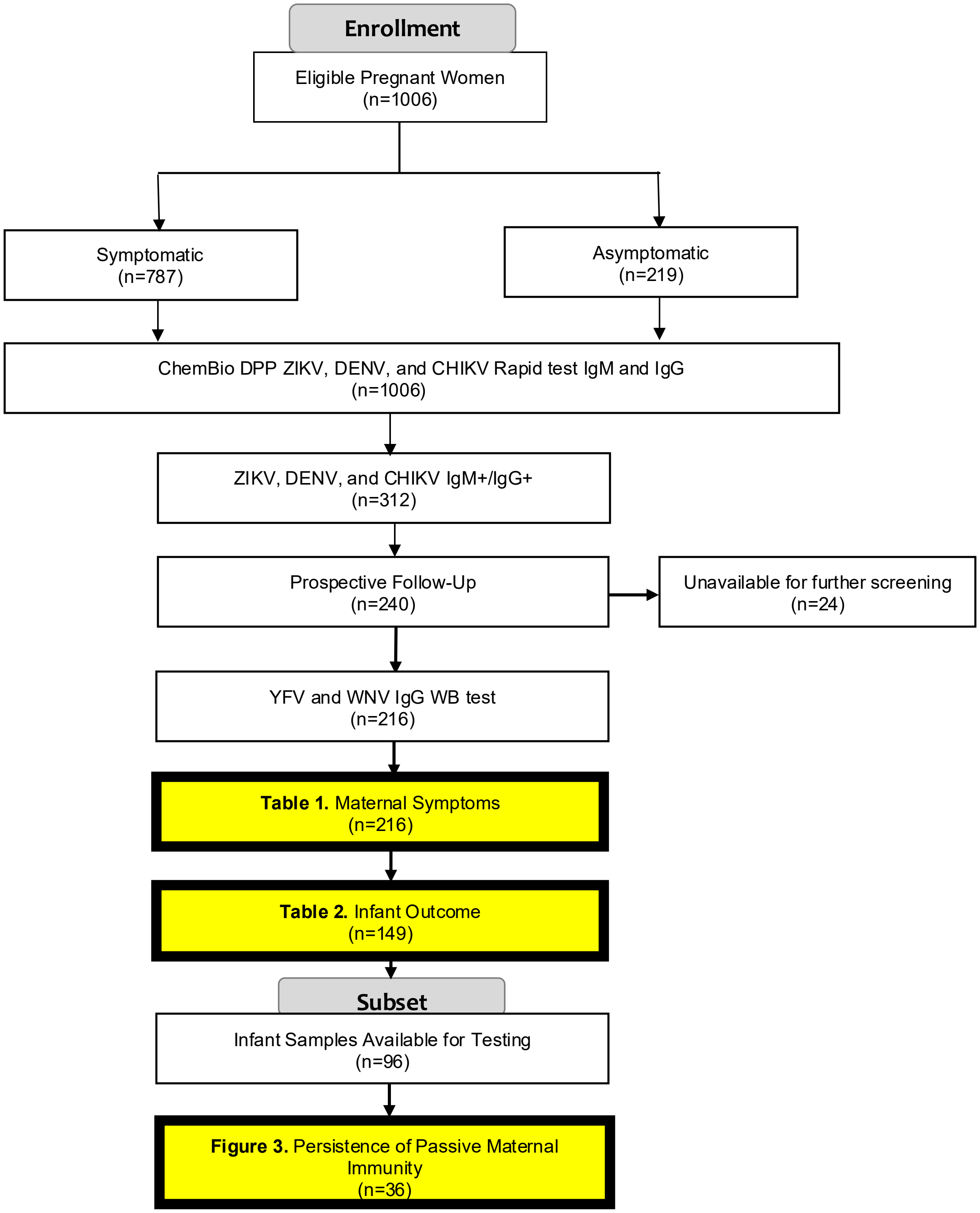


**Supplementary Figure 1. Study Flowchart**
Summary of participant enrollment, initial screening, and selection for serological analyses assessing YFV and WNV seroreactivity, maternal symptoms, infant outcomes, and passive maternal immunity.

| | Symptomatic | | Fisher's exact p-value | Logistic regression | |
|---|---|---|---|---|---|
| | No, # (%) | Yes, # (%) | | Odds ratio (95% CI) | p-value |
| YFV WB reactivity | | | | | |
| No | 42 (39.3) | 65 (60.8) | 0.114 | Ref | - |
| Yes | 31 (28.4) | 78 (71.6) | | **1.77 (1.21-2.61)** | **0.004** |
| ZIKV IgM reactivity | | | | | |
| No | 64 (35.2) | 118 (64.8) | 0.430 | Ref | - |
| Yes | 9 (26.5) | 25 (73.5) | | 1.49 (0.539-4.13) | 0.441 |
| DENV IgM reactivity | | | | | |
| No | 39 (29.1) | 95 (70.9) | 0.075 | Ref | - |
| Yes | 34 (41.5) | 48 (58.5) | | **0.514 (0.292-0.906)** | **0.021** |
| CHIKV IgM reactivity | | | | | |
| No | 37 (30.6) | 84 (69.4) | 0.311 | Ref | - |
| Yes | 36 (37.9) | 59 (62.1) | | 0.659 (0.377-1.15) | 0.143 |
| Site | | | | | |
| JUTH | 14 (14.6) | 82 (85.4) | **<0.001** | | |
| OLA | 59 (49.2) | 61 (50.8) | | | |
| Year screened | | | | | |
| 2019 | 26 (28.9) | 64 (71.1) | 0.220 | Ref | - |
| 2020 | 28 (41.8) | 39 (58.2) | | **0.546 (0.443-0.672)** | **<0.001** |
| 2021-22 | 19 (32.2) | 40 (67.8) | | **0.770 (0.732-0.811)** | **<0.001** |
| Age (years)* | 30 [26–34] | | | 0.997 (0.932-1.07) | 0.940 |
| Trimester | | | | | |
| 1st&2nd (1-27 wks) | 42 (36.2) | 74 (63.8) | 0.473 | | |
| 3rd (≥28 wks) | 31 (31.3) | 68 (68.7) | | | |

**Supplementary Table 1** Associations between YFV seroreactivity and maternal symptoms upon subsequent arbovirus infection among pregnant women in Jos, Nigeria. Statistical analyses include Fisher's exact test and logistic regression.

*Median [interquartile range] is shown for Age (years).

| | Abnormal infant outcome | | Fisher's exact p-value | Logistic regression | |
|---|---|---|---|---|---|
| | No, # (%) | Yes, # (%) | | Odds ratio (95% CI) | p-value |
| YFV WB reactivity | | | | | |
| No | 49 (75.4) | 16 (24.6) | 0.364 | Ref | - |
| Yes | 57 (67.9) | 27 (32.1) | | 1.29 (0.554-3.02) | 0.551 |
| ZIKV IgM reactivity | | | | | |
| No | 86 (71.1) | 35 (28.9) | 1.000 | Ref | - |
| Yes | 20 (71.4) | 8 (28.6) | | 1.01 (0.578-1.75) | 0.983 |
| DENV IgM reactivity | | | | | |
| No | 61 (79.1) | 26 (29.9) | 0.855 | Ref | - |
| Yes | 45 (72.6) | 17 (27.4) | | 1.37 (0.723-2.59) | 0.335 |
| CHIKV IgM reactivity | | | | | |
| No | 61 (79.2) | 16 (20.8) | **0.030** | Ref | - |
| Yes | 45 (62.5) | 27 (37.5) | | **2.50 (1.48-4.20)** | **0.001** |
| Site | | | | | |
| JUTH | 32 (68.1) | 15 (31.9) | 0.567 | | |
| OLA | 74 (72.6) | 28 (27.4) | | | |
| Year screened | | | | | |
| 2019 | 51 (66.2) | 26 (33.8) | 0.326 | | |
| 2020 | 26 (72.2) | 10 (27.8) | | | |
| 2021-22 | 29 (80.6) | 7 (19.4) | | | |
| Age (years) | 30 [26-33] | | | **1.07 (1.02-1.11)** | **0.004** |
| Trimester | | | | | |
| 1st&2nd (1-27 wks) | 60 (74.1) | 21 (25.9) | 0.468 | | |
| 3rd (≥28 wks) | 46 (67.7) | 22 (32.3) | | | |

**Supplementary Table 2.** Associations between YFV seroreactivity and abnormal infant outcomes upon subsequent arbovirus infection among pregnant women in Jos, Nigeria. Statistical analyses include Fisher's exact test and logistic regression.
*Median [interquartile range] is shown for Age (years).

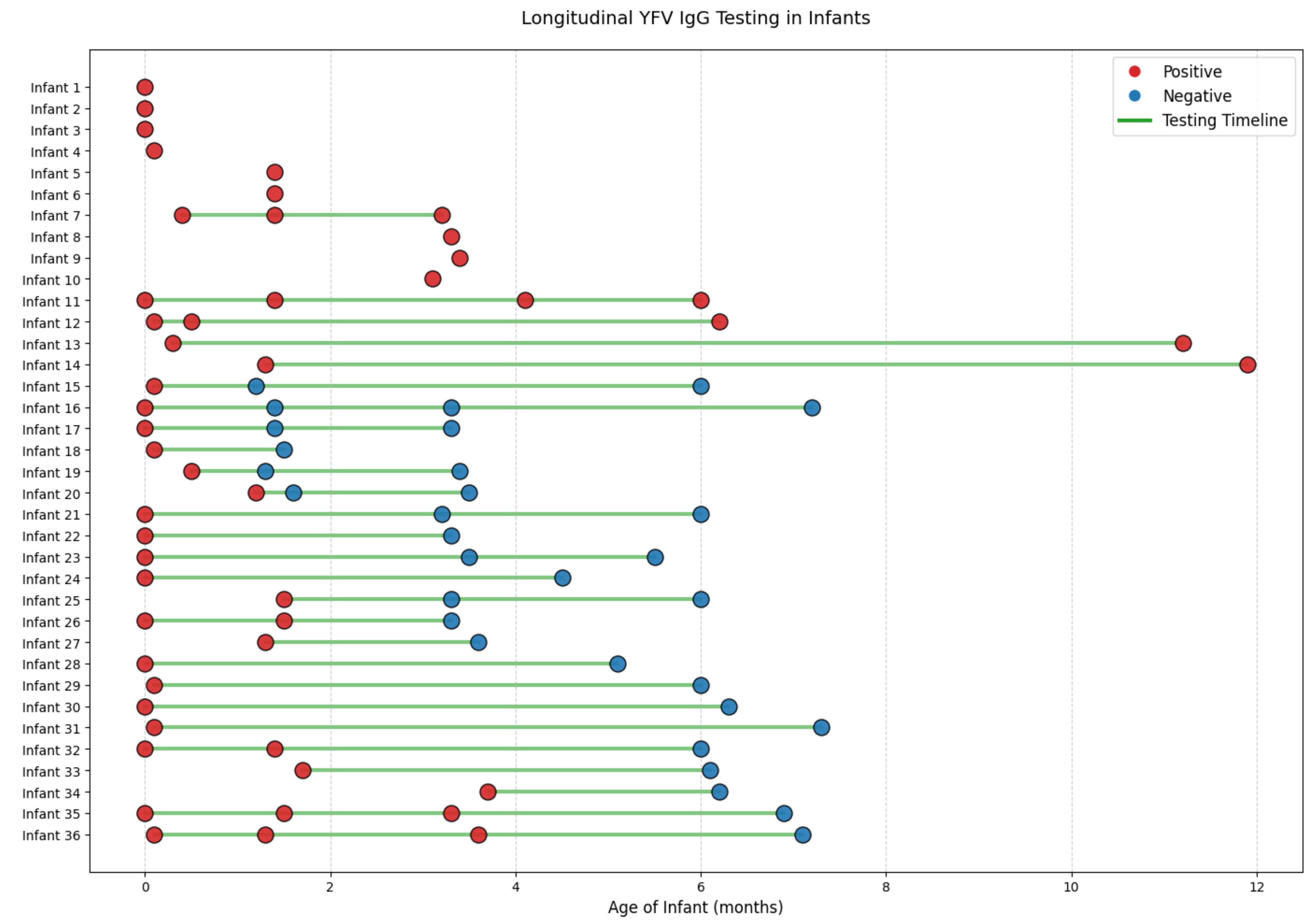


**Supplementary Figure 2. Longitudinal YFV IgG Testing in Infants**
Longitudinal follow-up of YFV IgG serostatus in infants over time. Each row represents an individual infant, with time points corresponding to the age at testing (in months). Red dots indicate positive YFV IgG results, blue dots indicate negative results, and gray dots indicate untested time points. Green lines represent the duration of follow-up for each infant.
